# A novel prediction model for immunotherapy induced pneumonitis prediction based on Chest CT and electronic health record

**DOI:** 10.1101/2024.10.14.24315487

**Authors:** Qing Lyu, Hongyu Yuan, Zhen Lin, Janardhana Ponnatapura, Christopher T Whitlow

**Affiliations:** Department of Radiology, Wake Forest University School of Medicine, Winston-Salem, NC, 27103, USA; Department of Biomedical Engineering, Wake Forest University School of Medicine, Winston-Salem, NC, 27103, USA

**Keywords:** Immunotherapy induced pneumonitis, immune checkpoint inhibitor, lung cancer, machine learning, deep learning, radiomics, computed tomography

## Abstract

Immune checkpoint inhibitors (ICIs) have been extensively used for the treatment of non-small cell lung cancer patients in recent years, providing a significant survival benefit. However, a major drawback of ICIs-related immunotherapy is the risk of developing post-surgical pneumonitis. In this study, we propose a deep learning-embedded, multi-modality prediction approach to assess whether patients will develop ICI-pneumonitis after receiving ICIs-based immunotherapy. This approach utilizes multi-modal data, including clinical data and pre-treatment lung screening computed tomography (CT) images. We extracted three types of features: 1) deep learning features from CT scans using a pretrained vision transformer, 2) radiomic features from CT scans using predefined radiomic algorithms, and 3) clinical features from patients’ clinical records. We then compared multiple machine learning algorithms for prediction based on these extracted features. Our results demonstrated a prediction accuracy of 0.823 and an area under the receiver operating characteristic curve of 0.895.

## Introduction

Lung cancer is the most frequently diagnosed cancer and the leading cause of cancer death worldwide^1^. Non-small cell lung cancer (NSCLC) is the most common subtype, accounting for approximately 85% of all lung cancer cases^2^. Recently, immunotherapy, particularly immune checkpoint inhibitors (ICIs), has demonstrated superior outcomes in the treatment of NSCLC, significantly improving overall survival compared to chemotherapy^3,4^. ICIs provoke immune reactions against cancer cells by blocking inhibitory receptors such as programmed cell death protein-1 (PD-1), programmed death-ligand 1 (PD-L1), and cytotoxic T-lymphocyte antigen 4 (CTLA-4)^5–7^. However, ICIs can also induce autoimmune reactions that are harmful to healthy tissues by disrupting normal immune system homeostasis, resulting in immune-related adverse events (irAEs)^8^. ICI-related pneumonitis (ICI-P) is a rare but life threatening irAE, with an overall incidence rate of 3-6% and a mortality rate of 22-33% for severe cases (grade 3-4)^9–11^. ICI-P can potentially cause significant morbidity, leading to the discontinuation of therapy and even mortality^8^. Clinical diagnosis of ICI-P is challenging due to its nonspecific symptoms and similarities to other pulmonary conditions^12^. There is no gold standard for clinical diagnosis, making it necessary to develop pretreatment ICI-P prediction methods.

Multiple studies have investigated the feasibility of predicting ICI-P before NSCLC patients receive immunotherapy. Risk factors such as tumor histologic type, ICIs selection, combination therapy, preexisting diseases (e.g. interstitial lung disease and extrathoracic metastasis), smoking history, and radiotherapy history are believed to influence the incidence rate of ICI-P^10,13–19^. Based on these findings, several prediction models have been developed. Jia *et al*. proposed a dynamic online hypertension nomogram to predict ICI-P for NSCLC patients^20^. Gong *et al*. developed a machine learning algorithm to identify and predict ICI-P based on eleven predictors^21^. Li and Xu both built risk assessment nomograms for ICI-P prediction^22,23^. Inaddition to using risk factors, researchers hypothesize that chest CT images contain discriminative information for ICI-P prediction. To validate this hypothesis, Colen *et al*. proposed a radiomics-based ICI-P prediction method, in extracting 1860 radiomic features from chest computed tomography (CT) images^24^. Mu *et al*. developed a radiomics nomogram to predict severe irAEs using fluorine-18 fluorodeoxyglucose positron emission tomography (PET) and CT images^25^. Cheng *et al*. created a deep learning embedded nomogram approach for ICI-P prediction, where a CT score, calculated from five radiology features extracted by a neural network, was input into a nomogram among with three other features for the final prediction^26^. Tan *et al*. constructed a multimodal deep learning model based on 3D CT images and clinical data, achieving an overall accuracy of 0.92 for ICI-P prediction through five-fold cross-validation^27^. Additionally, chest CT images can help discriminate different types of pneumonitis. Tohidinezhad *et al*. explored the feasibility of establishing a prediction model to differentiate ICI-P from other types of pneumonitis in NSCLC patients undergoing immunotherapy^28^. Mallio *et al*. utilized a deep learning algorithm based on chest CT images to distinguish coronavirus disease 2019 (COVID-19) pneumonia from ICI-P^29^.

Differing from existing studies, we propose a multimodal ICI-P prediction model utilizing three types of features: deep learning features, radiomic features, and clinical features. Deep learning features are extracted from a pretrained vision transformer (ViT), radiomic features are derived from predefined radiomic algorithms, and clinical features are obtained from patients’ clinical database. After feature selection, the selected features are input into the proposed ICI-P prediction model to predict the likelihood of ICI-P development following immunotherapy.

## Methodology

### Dataset

We collected data from 1,254 NSCLC patients who received immunotherapy between 2005 and 2021 at Wake Forest Baptist Medical Center. Among these, 51 patients developed ICI-P and were included in the experimental group for this study. Conversely, we randomly selected 41 patients who didn’t have ICI-P after immunotherapy to serve as the control group. Table 1 presents the demographic information of the patients. For each patient, we collected both the most recent chest CT scan taken before the first immunotherapy session and the corresponding clinical data.

**Table 1.** Patient demographic information.

|  | ICI-P | Control |
| --- | --- | --- |
| Amount | 51 | 41 |
| Gender | F: 21, M: 30 | F: 26, M: 15 |
| Age | 62.3±10.9 | 68.7±11.4 |
| BMI | 28.4±7.0 | 23.9±5.3 |

### Image processing

We first preprocessed the CT images to unify voxel resolution to 1×1×1 mm using nearest interpolation and cropped each image to a size of 512×512. At the same time, the pixel intensity was truncated to a range between −2100 and 100 Hounsfield units (HU). Lung segmentation was then implemented using three different networks: Lungmask^30^, nnUNet^31^, and Covid-19 MIScnn^32^. To achieve accurate lung segmentation, we developed a strategy that combines the results from these three networks. First, we created a union of the results from Lungmask, nnUNet, and Covid-19 MIScnn. Then, we manually checked each result from the previous step and corrected misclassified regions.

### Vision transformer

Given the huge success of transformers in natural language processing, ViT have recently been proposed as a competitive alternative to convolutional neural networks for various computer vision task, such as image classification, object detection, and semantic image segmentation^33^. In this study, we pretrained a ViT model to extract deep learning features from 3D CT images. A ViT-base model was pretrained with a hidden size of 768, 12 heads, and a depth of 12, following the strategy outlined by Niu *et al*.^34^.

During deep learning feature extraction, the CT images were divided into multiple 4×16×16 patches and fed into the pretrained ViT model. Each input patch produced a vector with a dimension of 768, and the summation of all vectors was used as the final set of deep learning features. For each patient, there were 768 deep learning features extracted.

### Radiomics

We utilized the Python Pyradiomics library for radiomic feature extraction^35^. To enhance the utilization of information within CT images, wavelet transform was applied before the feature extraction process. For each patient, there were 863 radiomic features extracted, categorized into 8 major categories: first order statistics, shape-based (2D), shape-based (3D), gray level co-occurrence matrix, gray level run length matrix, gray level size zone matrix, neighboring gray tone difference matrix, and gray level dependence matrix.

### Clinical data

Ten clinical features were extracted from patients’ electronic health record: 1) pack years, 2) age, 3) body mass index (BMI), 4) baseline oxygen dependence, 5) whether received surgery prior to immunotherapy, 6) whether received radiation prior to immunotherapy, 7) eastern cooperative oncology group performance status (ECOG PS) at the time of immunotherapy, 8) choice of immunotherapy, 9) whether immuno-oncology (IO) given concurrently with chemotherapy, and 10) total cycles of IO given. Since some clinical features are categorical and not represented numerically, we converted these features to one-hot encoding for prediction.

### Feature selection

After feature extraction, we obtained a total of 768 deep learning features, 863 radiomic features, and 10 clinical features. To remove redundant and non-relevant features, we performed feature selection using the Chi-square test and Student’s t-test. For deep learning and radiomic features, we retained only the 25 most significant features of each type. All 10 clinical features were kept. As a result, a total of 60 features were utilized for prediction after feature selection.

### Prediction model

We utilized the Python Pycaret library to establish ICI-P models. Ten different prediction models were compared, including logistic regression, K neighbors classifier, support vector machine, gradient boosting classifier, ada boost classifier, decision tree classifier, light gradient boosting machine, extra trees classifier, naïve bayes, and random forest classifier.

### Experimental details

We pretrained the ViT model on 2,000 chest CT scans collected by Wake Forest Baptist Medical Center between 2015 and 2021. The pretraining process was carried out for 100 epochs on an Nvidia A100 GPU. During pretraining, the original CT images were divided into multiple smaller patches of size 64 by 64 by 64. When combining features extracted from different approaches, we first normalized each feature to standardize feature values, mitigating potential issues caused by mismatched feature value magnitudes. Clinical features were processed thorugh either one-hot encoding or manual labeling to convert discrete features into a continuous domain. When using PyCaret for ICI-P prediction, three-fold cross validation was employed.

## Results

### Patient characteristics

A total of 92 NLCSC patients who received at least one cycle of immunotherapy at Wake Forest Baptist Medical Center were enrolled in this study. Among these patients, 51 developed ICI-P, with a median time of 149 days between the initial immunotherapy date and ICI-P diagnosis date. Of the patients who developed ICI-P, 45% received Pembrolizumab, 31% received Nivolumab, 14% received Durvalumab, 4% received Atezolizumab, and the remaining received a combination of immune checkpoint inhibitors. All ICI-P patients were diagnosed with pneumonitis of Grade 2 or higher: 44% had Grade 2, 44% had Grade 3, 7% had Grade 4, and 9% had Grand 5 pneumonitis. The 41 patients who didn’t develop ICI-P were used as the control group. Among them, 62% received Pembrolizumab, 17% received Nivolumab, 5% received Durvalumab, 14% received Atezolizumab, and 2% received Lenvatinib.

When conducting Chi-square test and Student’s t-test for clinical feature selection, we identified four features, including total cycles of IO given, pack years, BMI at diagnosis, and age, that showed significantly differences between ICI-P and control groups, as shown in Table 2. A nomogram was constructed as a quantitative method to predict the risk of ICI-P in NSCLC patients, as show in Figure 2.

**Table 2.**
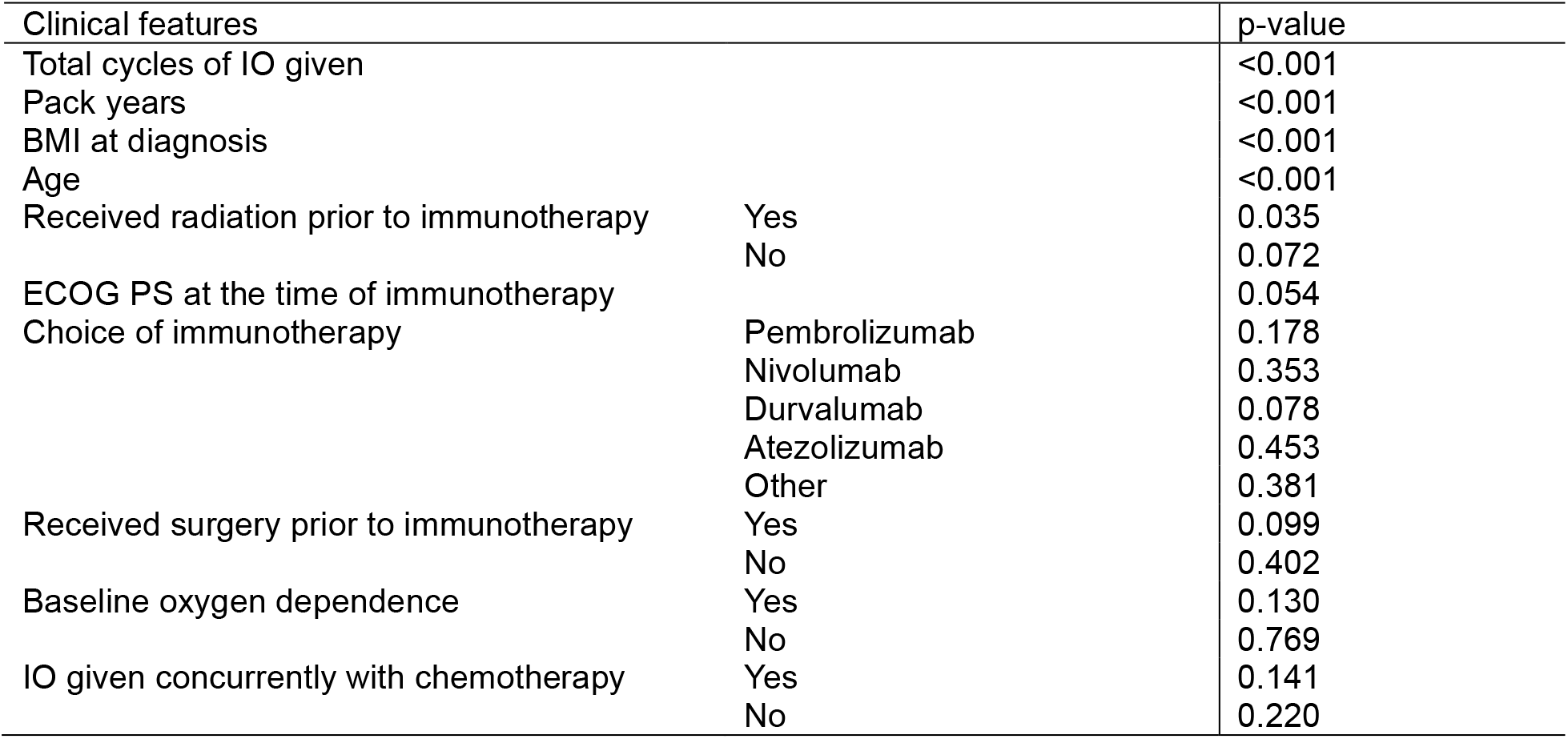
Identifying significant clinical features based on Chi-square test and Student’s t-test.

**Figure 1.**
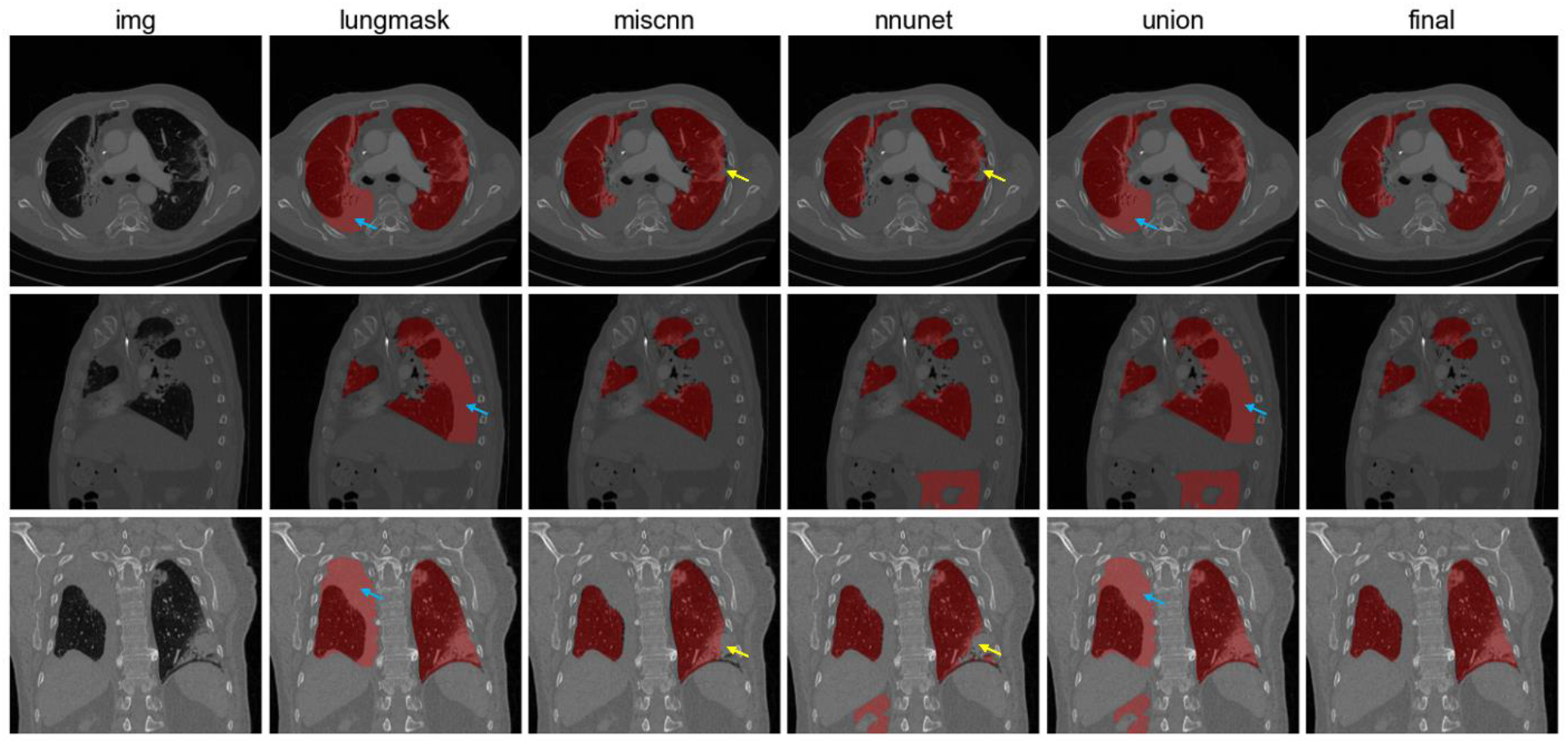
Lung segmentation results. From left to right: CT images, Lungmask segmentation, Covid-19 MIScnn, nnUNet, union results, and final results after manual correction. Blue arrows show false positive regions and yellow arrows demonstrate false negative regions.

**Figure 2.**
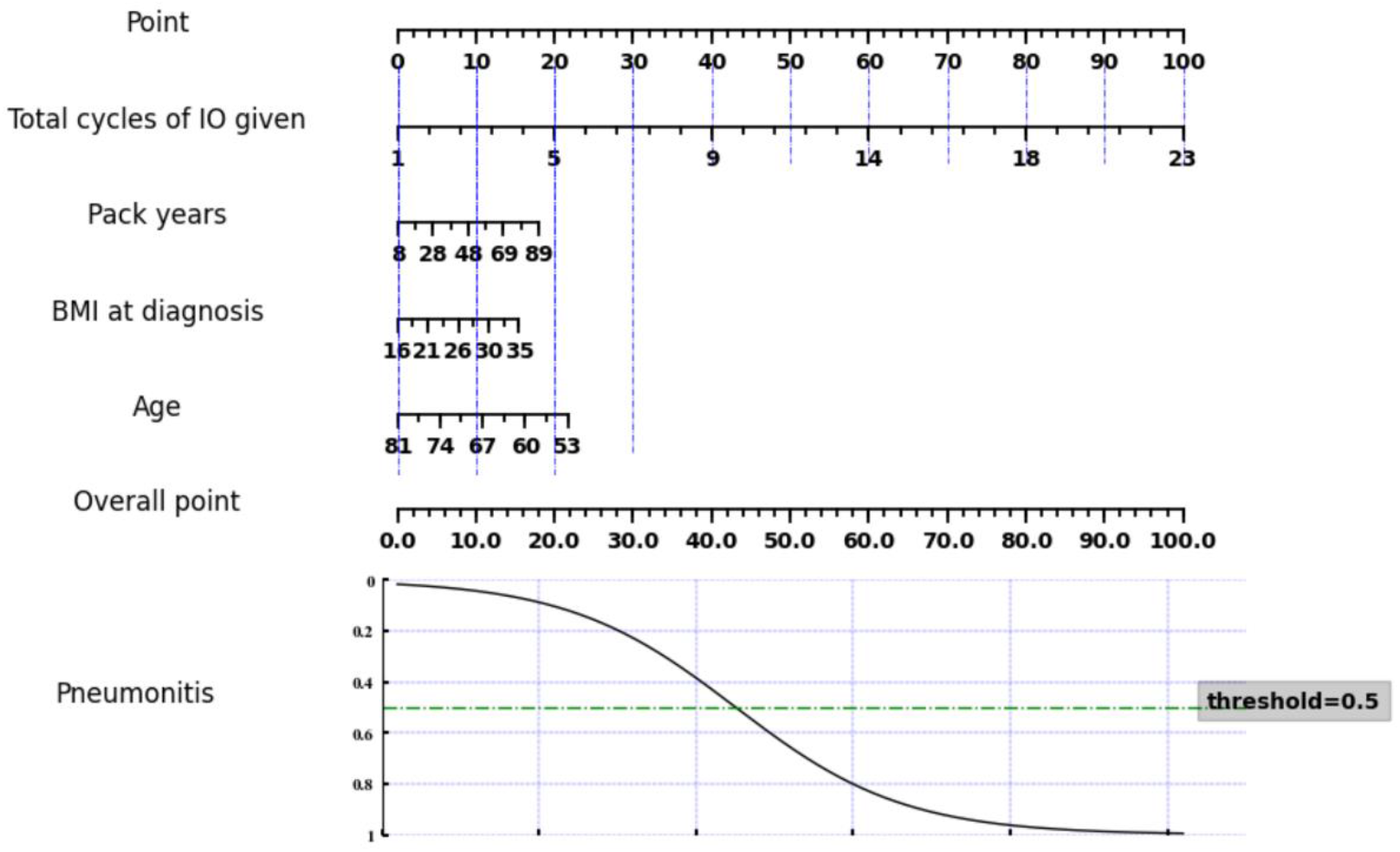
Nomogram of the ICI-P prediction model based on selected clinical features.

### ICI-pneumonitis prediction result based on all types of features

Metrics such as accuracy, area under the receiver operating characteristic curve (AUC), recall, precision, and F1 score were utilized to evaluate the performance of the ICI-P prediction models. Table 3 presents a comparison of different prediction models generated using PyCaret based on all selected multi-modal features. It can be found that logistic regression achieved the best performance in terms of accuracy, AUC, recall, and F1 score. Meanwhile, support vector machines achieved the highest precision.

**Table 3.** Comparison of different ICI-pneumonitis prediction models.

| Model | Accuracy | AUC | Recall | Precision | F1 |
| --- | --- | --- | --- | --- | --- |
| Logistic regression | **0.823** | **0.895** | **0.853** | 0.853 | **0.852** |
| K neighbor classifier | 0.809 | 0.892 | 0.787 | 0.882 | 0.830 |
| Support vector machines | 0.783 | 0.824 | 0.664 | **0.975** | 0.775 |
| Gradient boosting classifier | 0.746 | 0.818 | 0.789 | 0.791 | 0.788 |
| Ada boost classifier | 0.744 | 0.845 | 0.764 | 0.816 | 0.783 |
| Decision tree classifier | 0.709 | 0.722 | 0.662 | 0.811 | 0.725 |
| Light gradient boosting machine | 0.683 | 0.779 | 0.769 | 0.716 | 0.732 |
| Extra tree classifier | 0.683 | 0.711 | 0.811 | 0.714 | 0.757 |
| Naïve Bayes | 0.658 | 0.618 | 0.791 | 0.700 | 0.736 |
| Random forest | 0.631 | 0.726 | 0.767 | 0.672 | 0.714 |

### Ablation study

We conducted an ablation study to assess the contribution of each modality of features to the final prediction results and to demonstrate the effectiveness of the proposed method. A series of experiments were performed, building prediction models under four situations: using clinical features alone, deep learning features alone, radiomic features alone, and a combination of all features. As show in Table 4. The results indicate that using all three kinds of features together leads to higher accuracy and AUC scores compared to using each type of features individually. The only exception was the random forest model, which performed better when the prediction was based solely on clinical features.

**Table 4.** Comparison of multiple prediction models on different types of features.

| Model | accuracy |  |  |  | AUC |  |  |  |
| --- | --- | --- | --- | --- | --- | --- | --- | --- |
|  | Clinical | Radiomic | Deep | All | Clinical | Radiomic | Deep | All |
| LR | 0.802 | 0.721 | 0.711 | **0.823** | 0.835 | 0.723 | 0.757 | **0.895** |
| KNN | 0.790 | 0.691 | 0.655 | **0.809** | 0.826 | 0.712 | 0.671 | **0.892** |
| SVM | 0.723 | 0.677 | 0.688 | **0.783** | 0.794 | 0.692 | 0.692 | **0.824** |
| GBC | 0.727 | 0.661 | 0.644 | **0.746** | 0.792 | 0.712 | 0.677 | **0.818** |
| RF | **0.713** | 0.555 | 0.577 | 0.631 | **0.760** | 0.679 | 0.685 | 0.726 |

LR: linear regression, KNN: k neighbors classifier, SVM: support vector machines, GBC: gradient boosting classifier, RF: random forest.

## Discussion

Immune checkpoint inhibitor immunotherapy is a revolutionary treatment for NSCLC that leverages the body’s immune system to target and destroy cancer cells. ICIs significantly improve outcomes for NSCLC patients, leading to longer overall survival and durable responses for some patients compared to traditional chemotherapy^3,4^. However, a major side effect of ICI-related immunotherapy is the potential development of irAEs, particularly ICI-P, which, although rare, can be life-threatening. In this paper, we propose a multi-modal approach to predict the occurrence of ICI-P in patients undergoing ICI immunotherapy. The approach incorporates three types of features: clinical features from patients’ electronic health records and radiomic and deep learning features extracted from CT scans. Our study demonstrates that using all three types of features together yields the best predictive performance.

In our ablation study, we found that clinical features contributed more to the final prediction results compared to radiomic and deep learning features. This is expected, as clinical features are directly related to the patients’ health conditions and their treatment processes. In contrast, radiomic and deep learning features, extracted from CT scans, provide only implicit information about patients’ health status. Our experiments showed that the best prediction results were achieved when all three types of features were combined. This suggests that CT scans provide additional valuable information beyond what is available in the patients’ electronic health records, and this information can be effectively extracted using radiomic and deep learning algorithms.

Developing methods to predict ICI-P can lead to improved treatment outcomes. Accurate prediction of ICI-P allows for the early identification of patients who are at a higher risk of developing pneumonitis. This enables proactive monitoring, early intervention, and potentially modifying or discontinuing treatment to prevent severe outcomes. Predicting ICI-P risk can also help oncologists personalize treatment regimens. For patients at high risk, alternative therapies, dose adjustments, or combination strategies that lower the risk may be considered. In addition, predicting and managing ICI-P early can help prevent unplanned treatment interruptions, ensuring patients can continue their cancer therapy as planned and potentially improve overall outcomes.

## Data Availability

All data produced in the present study are available upon reasonable request to the authors

